# Mental health, longitudinal ART adherence, and viral suppression among adolescents and adults living with HIV in South Africa: a cohort study

**DOI:** 10.1101/2022.05.22.22275437

**Authors:** Andreas D. Haas, Raphael Lienhard, Christiane Didden, Morna Cornell, Naomi Folb, Tebatso M. G. Boshomane, Luisa Salazar-Vizcaya, Yann Ruffieux, Patience Nyakato, Anja E. Wettstein, Mpho Tlali, Mary-Ann Davies, Per von Groote, Milton Wainberg, Gary Maartens, John A. Joska

## Abstract

**Introduction:** Mental disorders are highly prevalent among people living with HIV and are associated with adverse HIV treatment outcomes. We assessed HIV treatment outcomes in patients with and without mental health diagnoses by sex and age.

**Methods:** Using hospital, outpatient and pharmacy claims and laboratory data from 2011 to 2020, we followed HIV-positive adolescents and adults aged ≥15 years who enrolled in a South African private sector HIV treatment programme. We performed a longitudinal trajectory analysis to identify patients with similar adherence patterns and examined associations between mental health diagnoses and adherence patterns using multinomial logistic regression. We examined non-adherence and viral non-suppression (VNS, viral load >400 copies/mL) in patients with and without mental health diagnoses by sex and age using mixed-effects Poisson regression models.

**Results:** 54,378 patients were followed for a median of 3.5 years (IQR 1.9-6.4), 20,743 (38%) of whom had received a mental health diagnosis. 90% of patients had HIV viral load <400 copies/mL, 73% had continuously high adherence, 13% had decreasing adherence, 6% had increasing adherence, and 7% were continuously non-adherent. Mental health diagnoses were associated with decreasing adherence (aRR 1.41, 95% CI 1.28-1.55), increasing adherence (aRR 1.59, 95% 1.41-1.79), and continuous non-adherence (aRR 2.02, 95% 1.81-2.25). The risk of VNS was increased among patients with organic mental disorders (aRR 1.55, 95% CI 1.22-1.96), substance use disorders (aRR 1.53 95% CI 1.19-1.97), serious mental disorders (aRR 1.30, 95% CI 1.09-1.54), and depression (aRR 1.19, 95% CI 1.10-1.28) compared to patients without mental health diagnoses. The risk of VNS was also increased among men (aRR 1.48, 95% CI: 1.31-1.67), adolescents (15-19 years, aRR 2.72, 95% CI 2.29-3.24) and young adults (20-24 years, aRR 2.29, 95% CI 1.83-2.85) compared to adults aged 25-34 years. Adolescents and young adults with and without mental health diagnoses had low viral suppression rates (≤66%); women aged 45 or older with mental health diagnoses had suppression rates of 92-95%.

**Conclusion:** Our study highlights the need for psychosocial interventions to improve HIV treatment outcomes, particularly for adolescents and young adults, and supports strengthening mental health services in paediatric and general HIV treatment programs.

## Introduction

South Africa has the largest HIV epidemic in the world, with approximately 7.8 million people living with HIV and over 5 million people receiving antiretroviral therapy (ART) [1]. Widespread access to ART has improved the life expectancy of people living with HIV [2], but the long-term effectiveness of ART depends on lifelong retention in HIV care, adequate ART adherence, and viral suppression [3,4].

Mental health and substance use disorders are highly prevalent among people living with HIV [5–8]. Studies have consistently identified higher rates of mental health and substance use disorders experienced by people living with HIV than those in the general population [9].

The co-occurrence of mental illness and HIV poses challenges to treating HIV. Mental disorders are associated with poor HIV treatment outcomes, including low adherence [7,10–12], poor virological outcomes [13–16], poor retention in HIV care [16,17] and increased mortality [16]. Estimates on associations between mental health and HIV treatment outcomes vary widely, likely due to methodological limitations of previous studies including cross-sectional study designs, small sample sizes, and the use of self-reported mental health and adherence measures.

Disparities in HIV treatment outcomes between age groups and between men and women are of concern. HIV treatment outcomes are typically worse in younger persons and men compared to older persons and women [18–22].

As South Africa is approaching the global HIV treatment target of 95% viral suppression among persons receiving ART [1], disaggregated data are needed to identify populations at the greatest risk of poor HIV treatment outcomes [23]. In this cohort study, we assessed HIV treatment outcomes in patients with and without mental health diagnoses by sex and age.

## Methods

### Study design

We followed a cohort of HIV-positive adolescents and adults who enrolled in the Aid for AIDS (AfA) HIV management programme from their first documented ART use (baseline) to the end of insurance coverage, death, or database closure, whichever occurred first. AfA has ethical approval to contribute data to the International epidemiology Databases to Evaluate AIDS (IeDEA) [24]. The Human Research Ethics Committee of the University of Cape Town, South Africa, and the Cantonal Ethics Committee Bern, Switzerland, authorised the analysis of the database. Beneficiaries of the medical insurance scheme or their guardians provided consent for their data to be used in research.

### Setting

The AfA programme is a private sector HIV management programme for insured people living with HIV in South Africa. Private medical practitioners and specialists treat HIV patients according to national treatment guidelines [25]. General practitioners, psychiatrists, psychologists, and private inpatient mental health facilities provide mental health care. Patients accessing HIV care in the public sector were not included in this study.

### Eligibility

HIV-positive adolescents and adults aged 15 years or older, receiving ART for at least six months, who had insurance coverage with a large South African medical insurance scheme between January 1, 2011, and June 30, 2020, and enrolled in the AfA programme were eligible for inclusion in this study. Individuals with missing sex or date of birth were excluded. Patients with less than one year of follow-up were excluded from the analysis of factors associated with adherence, those with less than three years of follow-up were excluded from adherence trajectory analysis, and those without viral load measurement were excluded from analysis of viral suppression (Figure S1).

### Data

We extracted demographic and laboratory data and reimbursement claims from the IeDEA database [24]. Pharmacy claims contained information on the active ingredients of drugs coded according to the Anatomical Therapeutic Chemical (ATC) classification system [26], the drug strengths, the dispensed amount, and the date of dispensing. Outpatient and hospitalisation claims contained International Classification of Diseases, 10th Revision (ICD-10) diagnoses [27]. Laboratory data contained HIV viral load and CD4 cell counts. Claims data were available from Jan 1, 2011, to Jun 30, 2020, and laboratory data from Jan 1, 2016 to June 30, 2020.

### Outcomes

We defined viral non-suppression (VNS) as an HIV viral load ≥400 copies/mL and explored thresholds of 100 copies/mL and 1000 copies/mL in sensitivity analyses. We assessed adherence based on pharmacy claims for antiretroviral medication (ATC codes J05AR, J05AG, J05AE, J05AJ, or J05AX). We calculated the duration of each claim by dividing the amount of active ingredient dispensed by the WHO-defined daily average maintenance dose for adults [28]. We assumed that patients who refilled their prescriptions early stockpiled unused drugs. We calculated patients’ continuous medication availability (CMA) in two steps [29,30]. First, we assigned the mean adherence value of an interval between two consecutive refills, or between the last refill and the end of the patient follow-up, to each day of the interval by dividing the number of days covered by sufficient drug supply during the interval and the number of days of the interval. Second, we split patients’ follow-up time into consecutive 1, 3, 6, and 12-month intervals and averaged daily mean adherence values over each interval. We defined non-adherence as CMA values below 80% and explored thresholds of 70% and 90% in sensitivity analyses.

### Exposures

Mental health was assessed based on ICD-10 diagnoses from outpatient and hospital claims. We considered diagnoses for any mental disorders in the ICD-10 range F00-F99. We categorised mental disorders into the following groups: organic mental disorders (ICD-10 codes F00-09), substance use disorders (F10-F19), serious mental disorders (F20-F29, and F31), depression (F32, F33, and F34.1), anxiety (F40-F48), or other mental disorders (F30, F34.0, F34.8, F34.9, F50-F99). In sensitivity analyses, we considered patients diagnosed if they had received at least two diagnoses on different dates. We grouped age into seven categories (15-19, 20-24, 25-34, 45-54, 55-64, and ≥65 years).

### Statistical analysis

Using summary statistics, we described patient characteristics by mental health status. To validate the CMA measure, we estimated the true-positive rate, false-positive rate, and area under the curve (AUC) of the CMA in 1, 3, 6, and 12 months before viral load testing for predicting VNS. We estimated these measures using receiver operating characteristic (ROC) regression models with probit link by maximum likelihood estimation.

We estimated unadjusted and adjusted risk ratios (RR) for factors associated with non-adherence and VNS using mixed-effects Poisson regression models with robust standard errors and a random intercept at patient level [31,32]. First, we estimated RRs for each group of mental health diagnoses, adjusting for age, sex, and years since baseline. Year since baseline was rounded to the next integer and modelled as a categorical variable. Mental health diagnoses, age, and the year since baseline were modelled as time-varying covariates. Second, we estimated RRs for each group of mental health diagnoses, adjusting for age, sex, year since baseline, and psychiatric comorbidity. Next, we adjusted RRs for associations between mental health diagnoses and VNS for age, sex, year since baseline and CMA. Finally, we estimated and plotted adjusted model predictions for outcomes at two years after baseline for patients with and without mental health diagnoses by age and sex. More details on statistical methods are given in the appendix (Text S1).

We performed a longitudinal trajectory analysis to identify persons with similar adherence trajectories using the R package kml [33]. The package implements a k-means expectation-maximization algorithm to cluster observations with homogeneous longitudinal trajectories into distinct groups. In the trajectory analysis, we modelled participants’ continuous three-monthly CMA scores over five years after baseline. We imputed missing CMA scores for participants with less than five years of follow-up based on participants’ trajectory means [33]. We ran the algorithm five times to identify two to six adherence groups and chose the optimal group size based on clinical relevance and Calinski-Harabatz’s and Ray-Turi’s criteria [33]. Based on visual inspection of plots of the mean CMA groups, we labelled the adherence patterns as ‘continuous high adherence’, ‘decreasing adherence’, ‘increasing adherence’, and ‘continuous non-adherence’.

Finally, we performed a multinomial logistic regression analysis to examine factors associated with adherence group affiliation. The regression model included a categorical variable for age, binary variables for sex and mental health diagnosis at baseline, and an interaction term between age and sex. Using predictive margins, we estimated and plotted the probability of group affiliation by sex, age, and mental health status.

Statistical analyses were done in Stata (Version 16) and R (R version 3.6.3).

## Results

### Characteristics of patients

We followed 54,378 HIV-positive adolescents and adults for a median duration of 3.5 years (IRQ 1.9-6.4). The median age of patients at baseline was 40.1 years (SD 9.9), and the majority were women (59%). At the end of follow-up, 38% of patients had been diagnosed with at least one mental disorder. Anxiety (26%) and depression (20%) were the most prevalent mental health diagnoses (Table 1). Mental health diagnoses were more prevalent in women (43%) than in men (32%). The prevalence of mental health diagnoses peaked in men (34%) and women (48%) aged 45-54 years (Table S1).

**Table 1:** Characteristics of patients by mental health status at the end of follow-up.

|  | **No mental health diagnosis** |  | **Mental health diagnosis** |  | **Total** |  |
| --- | --- | --- | --- | --- | --- | --- |
|  | N=33,635 (61.9) |  | N=20,743 (38.1) |  | N=54,378 (100.0) |  |
| Age, years |  |  |  |  |  |  |
| 15-19 | 828 | (2.5) | 385 | (1.9) | 1,213 | (2.2) |
| 20-24 | 698 | (2.1) | 329 | (1.6) | 1,027 | (1.9) |
| 25-34 | 8,524 | (25.3) | 5,498 | (26.5) | 14,022 | (25.8) |
| 35-44 | 12,474 | (37.1) | 8,259 | (39.8) | 20,733 | (38.1) |
| 45-54 | 8,117 | (24.1) | 4,898 | (23.6) | 13,015 | (23.9) |
| 55-64 | 2,720 | (8.1) | 1,267 | (6.1) | 3,987 | (7.3) |
| 65+ | 274 | (0.8) | 107 | (0.5) | 381 | (0.7) |
| Mean (SD) | 40.2 | (10.1) | 39.8 | (9.4) | 40.1 | (9.9) |
| Sex |  |  |  |  |  |  |
| Male | 15,219 | (45.2) | 7,131 | (34.4) | 22,350 | (41.1) |
| Female | 18,416 | (54.8) | 13,612 | (65.6) | 32,028 | (58.9) |
| ART regimen at baseline |  |  |  |  |  |  |
| NNRTI-based | 29,650 | (88.2) | 18,118 | (87.3) | 47,768 | (87.8) |
| II-based | 200 | (0.6) | 131 | (0.6) | 331 | (0.6) |
| PI-based | 3,785 | (11.3) | 2,494 | (12.0) | 6,279 | (11.5) |
| Follow-up time, years |  |  |  |  |  |  |
| Median (IQR) | 3.0 | (1.5-5.4) | 4.9 | (2.8-7.6) | 3.5 | (1.9-6.4) |
| Mental health diagnoses |  |  |  |  |  |  |
| Organic mental disorder | 0 | (0.0) | 488 | (2.4) | 488 | (0.9) |
| Substance use disorder | 0 | (0.0) | 429 | (2.1) | 429 | (0.8) |
| Serious mental disorder | 0 | (0.0) | 1,235 | (6.0) | 1,235 | (2.3) |
| Depression | 0 | (0.0) | 11,000 | (53.0) | 11,000 | (20.2) |
| Anxiety | 0 | (0.0) | 14,248 | (68.7) | 14,248 | (26.2) |
| Other mental disorders | 0 | (0.0) | 3,446 | (16.6) | 3,446 | (6.3) |

Data are the number of participants and percentages if not stated otherwise.
Abbreviations: SD=standard deviation, IQR=interquartile range, ART=antiretroviral therapy, NNRTI=non-nucleoside reverse transcriptase inhibitors, II=integrase inhibitor, PI=protease inhibitor

### Validation of CMA

CMA measured over 12 months before viral load testing had an AUC of 0.82 (95% CI 0.81 to 0.83) for predicting VNS. CMA measured over a shorter duration had a slightly lower AUC (Figure S2).

### Mental health and adherence

Figure 1 shows the estimated mean CMA in the second year after baseline in patients with and without mental health diagnosis by sex and age. Mean CMA ranged from 69% (95% CI 62-76) in men aged 20-24 who had received a mental health diagnosis to 95% (95% CI 93-97) in women ≥65 years who had not received a mental health diagnosis. Women with and without mental health diagnoses aged 45 years or older had a higher mean CMA than men (Figure S3). We observed no sex differences in younger age groups. In a model adjusted for mental health diagnosis of any mental disorder, age, sex, year since baseline, patients with a mental health diagnosis (aRR 1.21, 95% CI 1.18-1.25), men (aRR 1.25, 95% CI 1.16-1.34), and younger age groups were at increased risk of non-adherence (CMA <80%) (Table 2). In a model adjusted for age, sex, year since baseline, and psychiatric co-morbidity, organic mental disorders (aRR 1.17, 95% CI 1.00-1.38), substance use disorders (aRR1.41, 95% CI 1.24-1.62), depression (aRR 1.14, 95% CI 1.10-1.18), and anxiety (aRR 1.17, 95% CI 1.13-1.21) were associated with non-adherence (Table 2).

**Table 2:** Unadjusted and adjusted risk ratios for factors associated with non-adherence (CMA <80%).

|  | Unadjusted risk ratio<br>(95% CI) | Adjusted risk ratio<sup>a</sup><br>(95% CI) | Adjusted risk ratio<sup>b</sup><br>(95% CI) |
| --- | --- | --- | --- |
| Mental health diagnoses |  |  |  |
| No mental health diagnosis | 1.00 | 1.00 | 1.00 |
| Organic mental disorder | 1.42 (1.21-1.66) | 1.31 (1.12-1.54) | 1.17 (1.00-1.38) |
| Substance use disorder | 1.87 (1.64-2.13) | 1.60 (1.41-1.83) | 1.41 (1.24-1.62) |
| Serious mental disorder | 1.32 (1.19-1.46) | 1.23 (1.11-1.36) | 1.06 (0.95-1.18) |
| Depression | 1.32 (1.28-1.37) | 1.21 (1.17-1.25) | 1.14 (1.10-1.18) |
| Anxiety | 1.33 (1.29-1.37) | 1.22 (1.18-1.26) | 1.17 (1.13-1.21) |
| Other mental disorders | 1.23 (1.15-1.31) | 1.10 (1.03-1.17) | 1.02 (0.96-1.09) |
| Any mental disorder | 1.33 (1.29-1.37) | 1.21 (1.18-1.25) |  |
| Age, y |  |  |  |
| 15-19 | 1.38 (1.27-1.49) | 1.35 (1.24-1.46) | 1.36 (1.25-1.48) |
| 20-24 | 1.29 (1.19-1.40) | 1.40 (1.27-1.54) | 1.40 (1.27-1.54) |
| 25-34 | 1.00 | 1.00 | 1.00 |
| 35-44 | 0.79 (0.76-0.81) | 0.72 (0.69-0.74) | 0.72 (0.69-0.74) |
| 45-54 | 0.63 (0.61-0.66) | 0.55 (0.52-0.57) | 0.55 (0.52-0.57) |
| 55-64 | 0.66 (0.62-0.70) | 0.54 (0.51-0.57) | 0.54 (0.51-0.57) |
| 65+ | 0.48 (0.39-0.58) | 0.37 (0.31-0.45) | 0.37 (0.31-0.46) |
| Sex |  |  |  |
| Male | 1.11 (1.07-1.14) | 1.25 (1.16-1.34) | 1.25 (1.16-1.34) |
| Female | 1.00 | 1.00 | 1.00 |

Abbreviations: CMA=cumulative medication availability, CI=confidence interval
<sup>a</sup> Risk ratios for each group of mental health diagnoses were adjusted for years since baseline, age and sex.<sup>b</sup> Risk ratios adjusted for years since baseline, age, sex, organic mental disorders, substance use disorders, serious mental disorders, depression, anxiety and other mental disorders.

**Figure 1.**
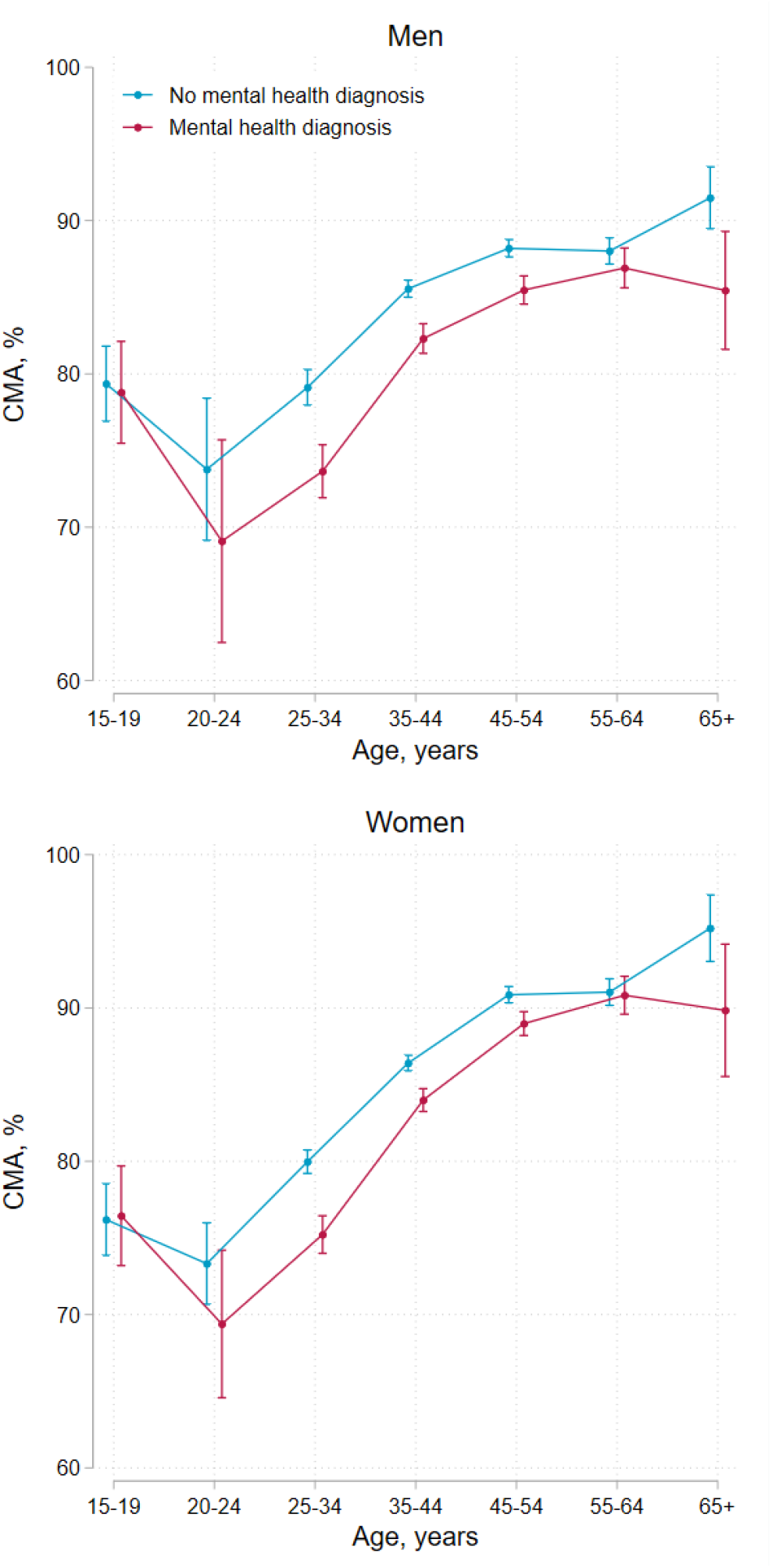
Cumulative medication availability (CMA) in the second year after baseline comparing patients with and without mental health diagnoses by age group and sex. Error bars represent 95% confidence intervals for means and proportions. N=48,645

In the adherence trajectory analysis, we identified four distinct longitudinal adherence trajectories. The estimated mean CMA for each group is shown in Figure 3A. Most patients (73%, 23,686/32,254) had continuously high adherence, 13% (4,152) had decreasing adherence, 6% (2,073) had increasing adherence, and 7% (2,343) were continuously non-adherent. Patients who had received a mental health diagnosis at baseline were more likely to have decreasing adherence (aRR 1.41, 95% CI 1.28-1.55), increasing adherence (aRR 1.59, 95% 1.41-1.79), or continuous non-adherence (aRR 2.02, 95% 1.81-2.25) compared to patients who had not received a mental health diagnosis. Young age was the strongest predictor of suboptimal adherence patterns (Table 4).

**Table 3:**
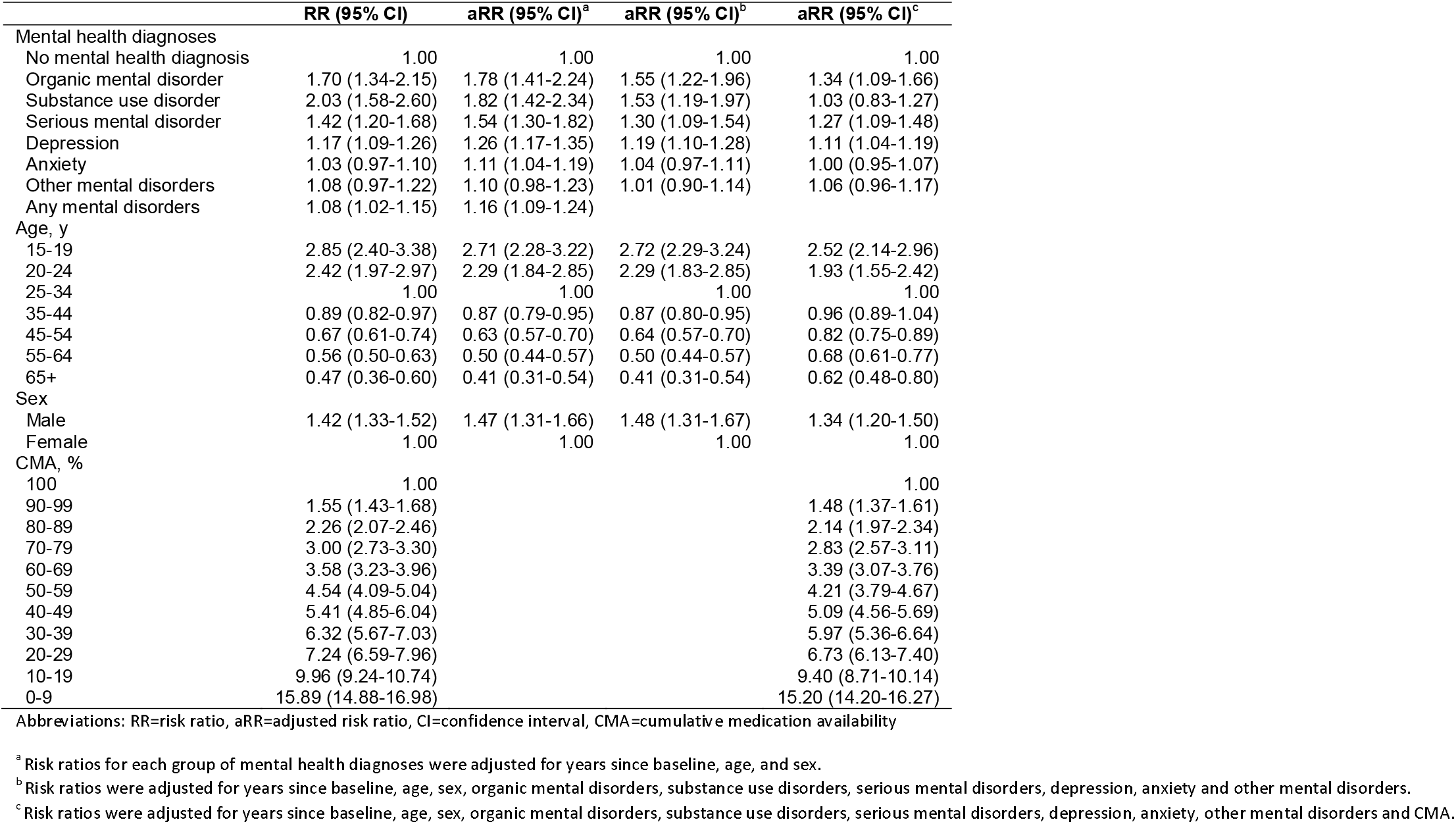
Unadjusted and adjusted risk ratios for factors associated with viral non-suppression (viral load ≥400 copies/mL)

**Table 4:**
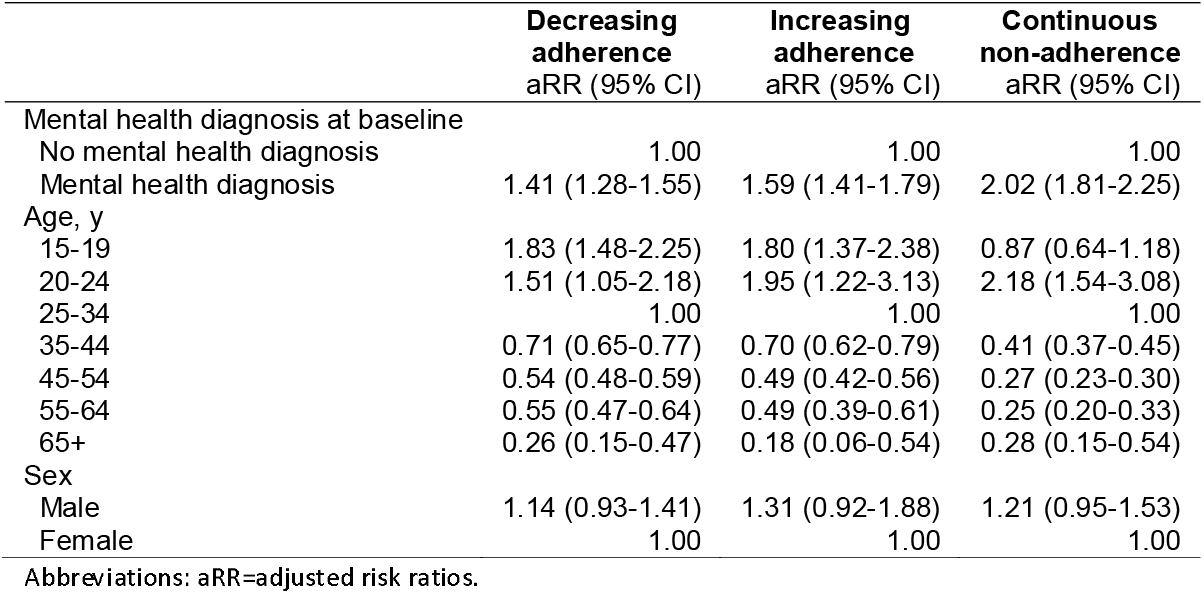
Risk ratios for factors associated with decreasing adherence, increasing adherence, or continuous non-adherence compared to continuously high adherence.

|  | **Decreasing**<br>**adherence**<br>**aRR (95% CI)** | **Increasing**<br>**adherence**<br>**aRR (95% CI)** | **Continuous**<br>**non-adherence**<br>**aRR (95% CI)** |
| --- | --- | --- | --- |
| Mental health diagnosis at baseline |  |  |  |
| No mental health diagnosis | 1.00 | 1.00 | 1.00 |
| Mental health diagnosis | 1.41 (1.28-1.55) | 1.59 (1.41-1.79) | 2.02 (1.81-2.25) |
| Age, y |  |  |  |
| 15-19 | 1.83 (1.48-2.25) | 1.80 (1.37-2.38) | 0.87 (0.64-1.18) |
| 20-24 | 1.51 (1.05-2.18) | 1.95 (1.22-3.13) | 2.18 (1.54-3.08) |
| 25-34 | 1.00 | 1.00 | 1.00 |
| 35-44 | 0.71 (0.65-0.77) | 0.70 (0.62-0.79) | 0.41 (0.37-0.45) |
| 45-54 | 0.54 (0.48-0.59) | 0.49 (0.42-0.56) | 0.27 (0.23-0.30) |
| 55-64 | 0.55 (0.47-0.64) | 0.49 (0.39-0.61) | 0.25 (0.20-0.33) |
| 65+ | 0.26 (0.15-0.47) | 0.18 (0.06-0.54) | 0.28 (0.15-0.54) |
| Sex |  |  |  |
| Male | 1.14 (0.93-1.41) | 1.31 (0.92-1.88) | 1.21 (0.95-1.53) |
| Female | 1.00 | 1.00 | 1.00 |

Abbreviations: aRR=adjusted risk ratios.

**Figure 2.**
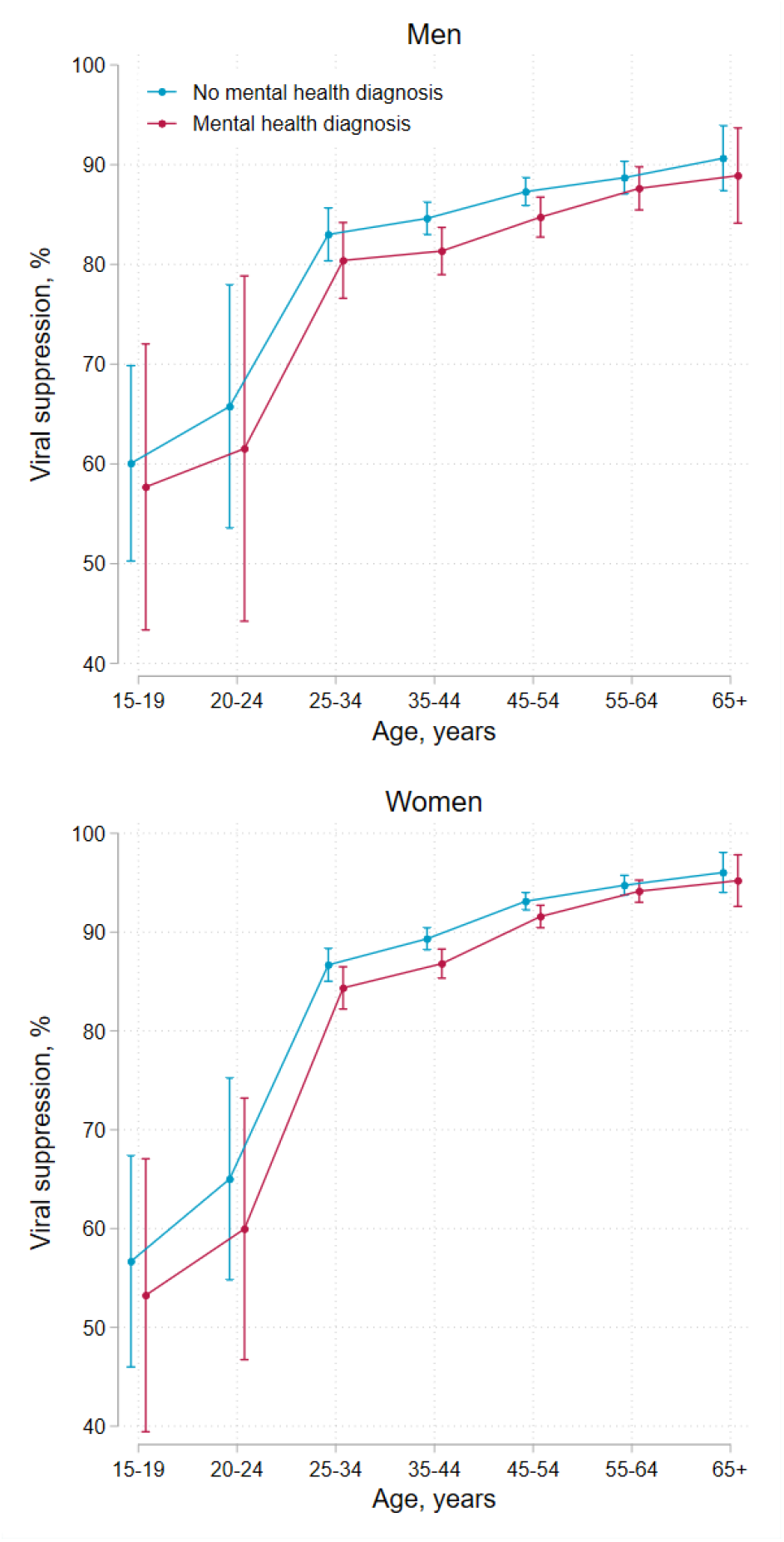
Viral suppression (viral load <400 copies/mL) at 2 years after baseline comparing patients with and without mental health diagnoses by age and sex. Error bars represent 95% confidence intervals for means and proportions. N=28,785

**Figure 3.**
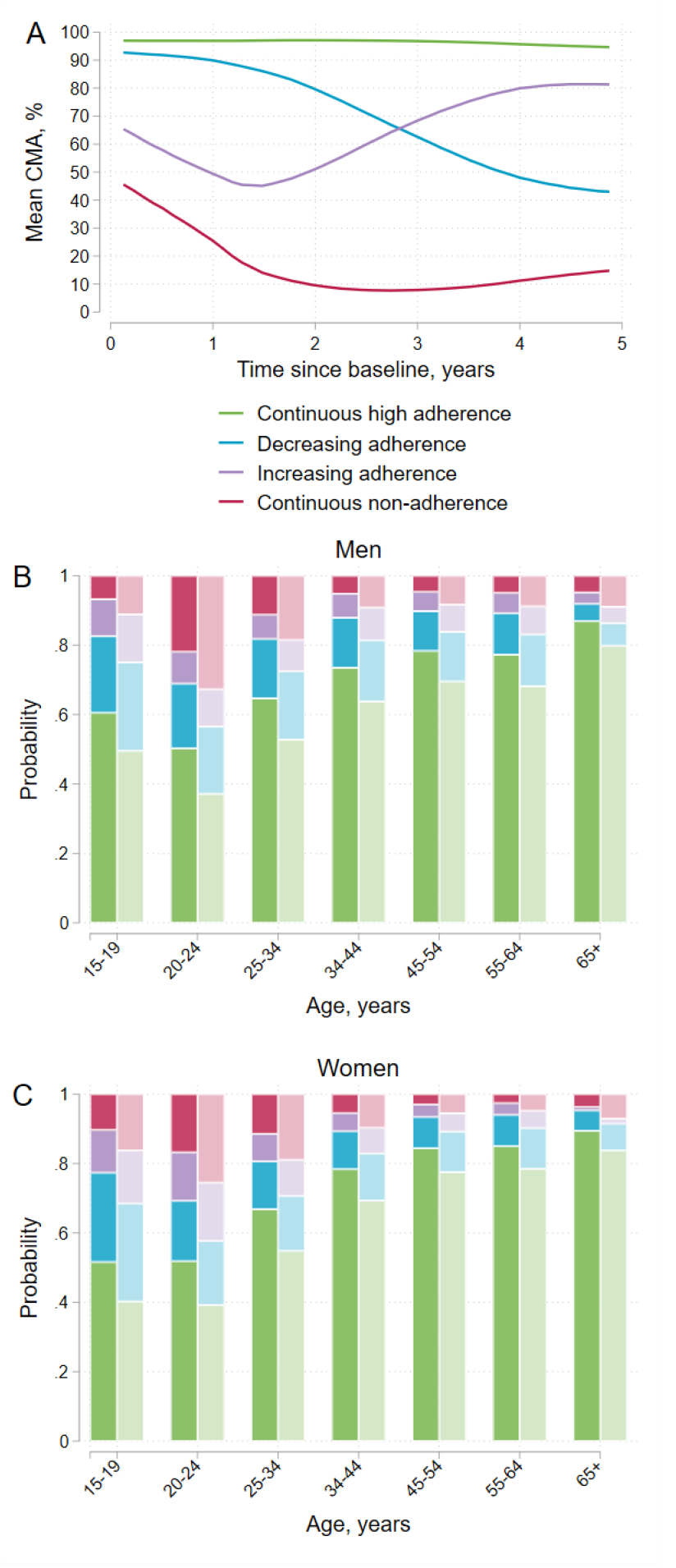
Adherence trajectories and the probability of being in each adherence group by sex, age and mental health diagnosis at baseline. Panel A shows the mean cumulative medication adherence (CMA) for the four groups identified in the longitudinal adherence trajectory analysis. Panels B and C show the probability of being in each group by sex, age, and mental health diagnosis at baseline. Darker colours represent persons who had not received a mental health diagnosis (ICD10 F00-F99) before or at baseline and lighter colours those who had received a mental health diagnosis. Participants with at least three years of follow-up were included in this analysis (N=32,254).

Figure 3B-C shows the predicted probabilities of being in each adherence group by sex, age, and mental health diagnoses at baseline. Suboptimal adherence patterns were more prevalent in younger age groups and patients with mental health diagnoses than in older age groups and those without mental health diagnoses. For example, men aged 20-24 years with mental health diagnoses had the highest risk of being continuously non-adherent (33%, 95% CI 20-46), whereas women aged 55-64 years without mental health diagnoses had the lowest risk (3%, 95% CI 2-4). Table S2 shows 95% CIs for predicted probabilities.

### Mental health and viral suppression

Overall, 90% (71,433/79,463) of the recorded viral load measurements were below the viral load threshold of 400 copies/mL. Figure 2 shows viral suppression (viral load <400 copies/mL) rates two years after baseline by sex, age, and mental health status. Viral suppression rates ranged from 53% (95% CI 39-67) in female adolescents aged 15-19 years with mental health diagnoses to 96% (95% CI 94-98) in women aged 65 years or older without mental health diagnoses. Viral suppression rates increased with increasing age and were lower in patients with mental health diagnoses. Women older than 25 years had higher suppression rates than men; adolescent girls and young women under 25 years had lower suppression rates than their male counterparts (Figure S4). Numerical values for viral suppression rates by sex, age group, and mental health status at two years after baseline for all three viral load thresholds (100, 400, and 1000 copies/mL) are shown in Table S3. In all adjusted models, male sex and younger age were strongly associated with VNS (Table 3). In models adjusted for age, sex, and years since baseline, VNS was associated with organic mental disorders (aRR 1.78, 95% CI 1.41-2.24), substance use disorders (aRR 1.82 95% CI 1.42-2.34), serious mental disorders (aRR 1.54, 95% CI 1.30-1.82), depression (aRR 1.26, 95% CI 1.17-1.35), anxiety (aRR 1.11, 95% CI 1.04-1.19), and other mental disorders (aRR 1.10, 95% CI 0.98-1.23). In models adjusted for age, sex, and psychiatric comorbidity, associations between all mental health diagnoses and VNS were attenuated; diagnoses of anxiety or other mental disorders were no longer associated (Table 3). The association between substance use disorders and VNS was fully mediated by CMA. Associations between other mental health diagnoses and VNS were partially mediated by CMA (Table 3).

### Sensitivity analyses

Conclusions were robust in sensitivity analyses with different thresholds for non-adherence (CMA<70% and <80%) and VNS (viral load >100 copies/mL and >1000 copies/mL) or when using an alternative exposure definition requiring patients to be diagnosed at least twice on different dates (Tables S4, Table S5).

## Discussion

In this cohort of privately insured adolescents and adults living with HIV, mental health diagnoses, younger age, and sex were associated with unfavourable adherence patterns and VNS. In patients with depression and anxiety, the increase in the risk of adverse HIV treatment outcomes was modest. In contrast, patients with serious mental disorders, substance use disorders, and organic mental disorders were at substantially higher risk of VNS and non-adherence compared to those without a mental health diagnosis. Adolescents and young adults with and without mental health diagnoses had low viral suppression rates, whereas older adults generally had high suppression rates, even those with mental health diagnoses. Male sex was associated with both increased risk of non-adherence (particularly young adults aged 20-24 years with mental illness) and with VNS.

Our results highlight the large burden of mental illness among adolescents and adults living with HIV in South Africa and underline the need to integrate mental health care into HIV treatment programs. Over a median follow-up duration of 3.5 years, 38% of the patients enrolled in this study had been diagnosed with a mental disorder; 20% had been diagnosed with depression and 26% with anxiety. The similarity between these figures and the estimated prevalence of depression (15%) and anxiety (23%) among people living with HIV in low- and middle-income settings suggests high rates of ascertainment of these disorders in this private sector program [5,6]. By contrast, the low proportion of patients diagnosed with substance use disorders likely reflects under-ascertainment of prevalent alcohol and substance use disorders [7,34]. Importantly, conclusions regarding access to mental health care cannot be generalised to patients accessing public sector HIV care programs. In a previous study, we showed that, in South Africa’s public sector, rates of ascertainment and treatment of mental disorders are much lower than in the private sector [35]. Interventions aimed at addressing the high burden of mental illness amongst people living with HIV are needed in both the public and private sectors in South Africa. Routine screening for mental health and substance use disorders [36,37] and task sharing of mental health counselling from specialised to non-specialised health workers or trained laypersons are promising approaches for integrating mental health services in primary care HIV care programs in low- and middle-income countries [38–42].

Our study confirms and extends previous findings on associations between mental health conditions and HIV treatment outcomes. In line with previous systematic reviews of studies on adults, we found depression and anxiety disorders associated with non-adherence. In a meta-analysis of 11 studies from Sub-Saharan Africa, people living with HIV and depression or depressive symptoms had 55% lower odds of achieving optimal ART adherence than those without depression or depressive symptoms [7]. These results were confirmed in a meta-analysis of 111 studies conducted in low-, middle- and high-income countries [11]. In another meta-analysis, anxiety symptoms were associated with 59% higher odds of suboptimal ART adherence [10]. Our study extends previous work by demonstrating that less prevalent but more serious mental disorders such as bipolar disorder, or schizophrenia are more strongly associated with non-adherence than common mental disorders. The involvement of significant others as treatment partners or directly observed therapy are recommended strategies to improve adherence in persons with serious mental illness [43].

Although patients diagnosed with a mental disorder were at an increased relative risk of VNS, the vast majority of them had good adherence and achieved viral suppression. Patients with mental illness should be assessed individually without implicit bias regarding their medication adherence [43]. Our data suggest that most patients with a mental illness have a good understanding of lifelong ART and do not need additional adherence support. Therefore, we believe that patients with mental illness should not per se be considered ineligible for differentiated ART delivery models for clinically stable patients [44,45]. While some mental health patients may benefit from closer monitoring and mental health interventions addressing mental health and adherence challenges [46], patients with well-controlled mental illness who are established on ART and adherent should have equal access to differentiated service delivery models [45].

Consistent with previous reports [18,19], adolescents and young adults had poorer HIV treatment outcomes than older adults. In our study, only about 40-60% of patients under 25 years had continuously high adherence. The proportion of patients with decreasing adherence was highest in adolescents aged 15 to 19 years. This decrease in adherence might reflect challenges related to transitioning from paediatric to adolescent care [47]. The risk of continuous non-adherence was highest in the young adults aged 20-24 years, peaking at 33% in young men with mental health diagnoses. In adolescents aged 15-19 years old, the risk of continuous non-adherence was 7-16%. Continuous non-adherence reflects long treatment interruption or discontinuation of ART. A multi-cohort study from South African public sector HIV treatment programs reported much higher rates of loss to follow-up (>60% at 2 years after ART initiation) in this age group [48]. Poor HIV treatment outcomes in adolescents and young adults highlight the need for interventions to improve care outcomes in this age group. A recent meta-analysis found that psychosocial interventions for adolescents and young people living with HIV showed small-to-moderate effects on adherence and viral load [49]. Scale-up of successful interventions to improve HIV outcomes of young people living with HIV should be a priority.

Strengths of this study include the large sample size, allowing for disaggregated analyses of common and less prevalent serious mental disorders by sex and age, and the availability of mental health diagnoses from primary, secondary, and tertiary care. Most previous studies relied on brief screening tools [7,10,11] that usually have a high false-positive rate and low positive predictive value for mental health diagnoses [50]. Further strengths of our study include the use of an objective validated adherence measure, the longitudinal study design, and the novel analytic methods used to examine longitudinal adherence patterns.

Our findings have to be considered in light of the following limitations. First, we classified patients’ mental health status based on ICD-10 diagnoses from reimbursement claims and thus missed patients with undiagnosed mental disorders. Patients with mild forms of mental disorders might be less likely to be diagnosed and those with more severe mental illness might be over-represented in our sample of mental health patients. A further limitation of administrative data is the possibility of miscoding of ICD-10 diagnoses. Nevertheless, mental health diagnoses from administrative data generally have a high positive predictive value for research diagnoses [51], and our conclusions held true when we considered only repeated mental health diagnoses in sensitivity analyses. Second, we used a pharmacy claim-based adherence measure that may overestimate adherence if patients do not take all collected medication, or underestimate adherence if drugs are obtained without documentation or from other sources (e.g., public sector clinics). Despite these limitations, objective pharmacy claim-based measures are considered more reliable than self-reported adherence measures [52] used in most previous studies [7,10,11]. In addition, the high accuracy of CMA for predicting HIV viral load validates our adherence measure. Third, because laboratory data were only available for the years 2016 to 2020, we could only include about half of the patients in analyses of viral suppression and VNS. Fourth, our study is not representative of the general population of people living with HIV in South Africa. We analysed data from a private sector HIV programme for employed and insured persons. Therefore, our findings cannot be generalised to persons accessing HIV care in the public sector.

## Conclusions

Our study confirms high rates of mental illness amongst people living with HIV in South Africa. Mental illness, particularly the less prevalent but potentially more serious mental disorders such as bipolar disorder and schizophrenia, young age and male sex were associated with poor treatment outcomes. However, the vast majority of patients with mental illness had good adherence and achieved viral suppression. The findings highlight the need for psychosocial interventions to improve HIV treatment outcomes, particularly for adolescents and young adults, and support strengthening mental health care in general and paediatric HIV treatment programs. Patients with mental illness who are established on ART and adherent to their medication should have equal access to differentiated service delivery models.

## Data Availability

Data were obtained from the IeDEA-SA. Data cannot be made available online because of legal and ethical restrictions. To request data, readers may contact IeDEA-SA for consideration by filling out the online form available at https://www.iedea-sa.org/contact-us/. Statistical code is available under https://github.com/AndreasDHaas/MH-CMA-VNS.

## Authors’ contributions

AH and RL conceived the study and wrote the first draft of the study protocol, which was revised by JJ, GM, CD, and YR. All authors reviewed and approved the final version of the study protocol. AH performed statistical analysis. CD, LSV, PN and YR advised on statistical methods. All authors contributed to interpretation of results. AH and RL wrote the first draft of the manuscript, which was revised by JJ, GM, MC, AW, YR, NF and LSV. All authors contributed to the final version of the manuscript and approved it for submission.

## Disclaimer

The content is solely the responsibility of the authors and does not necessarily represent the official views of the National Institutes of Health.

## Supplementary appendix

**Figure S1:**
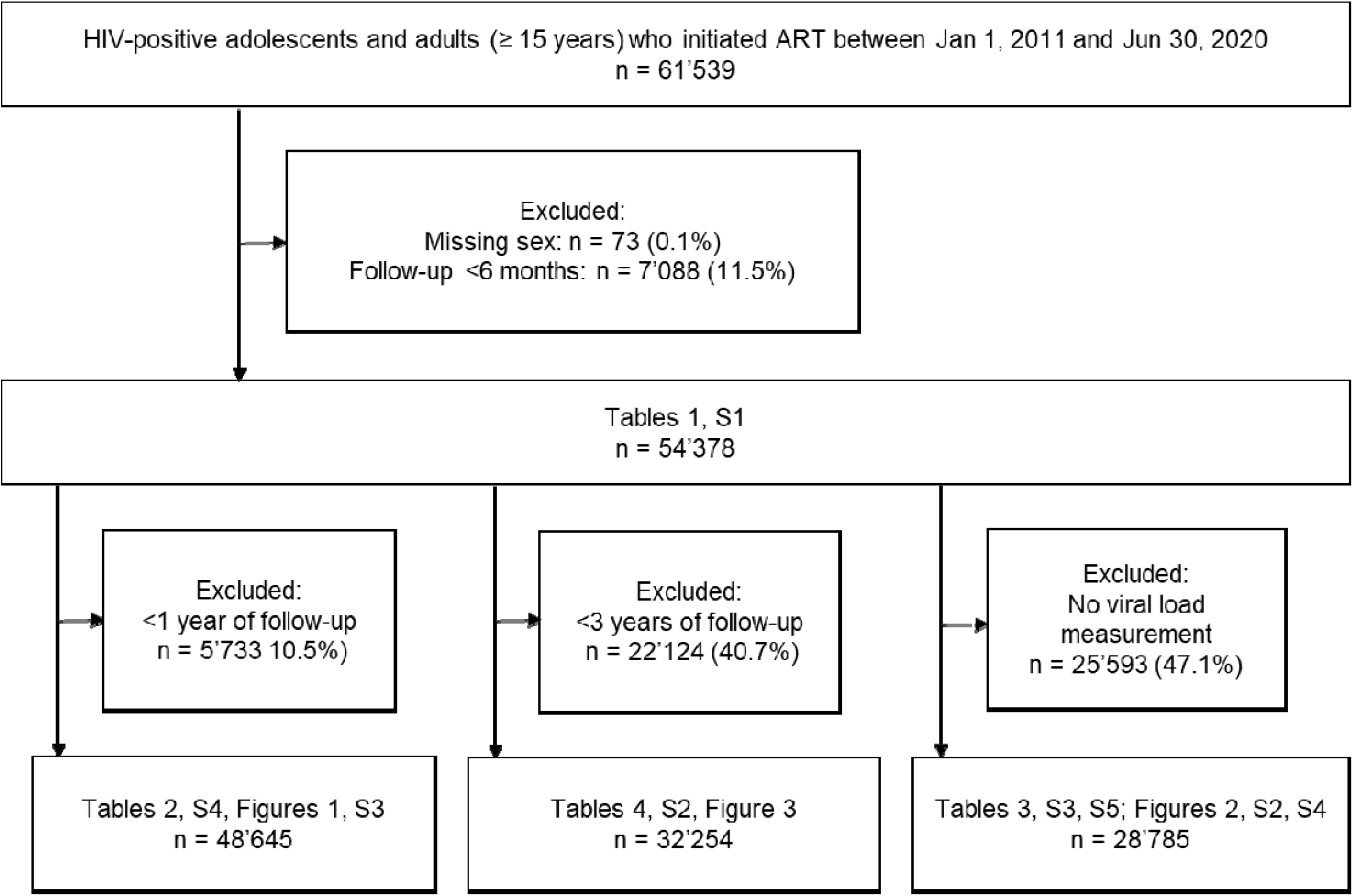
Flow diagram of eligibility of patients for analysis.

**Figure S2.**
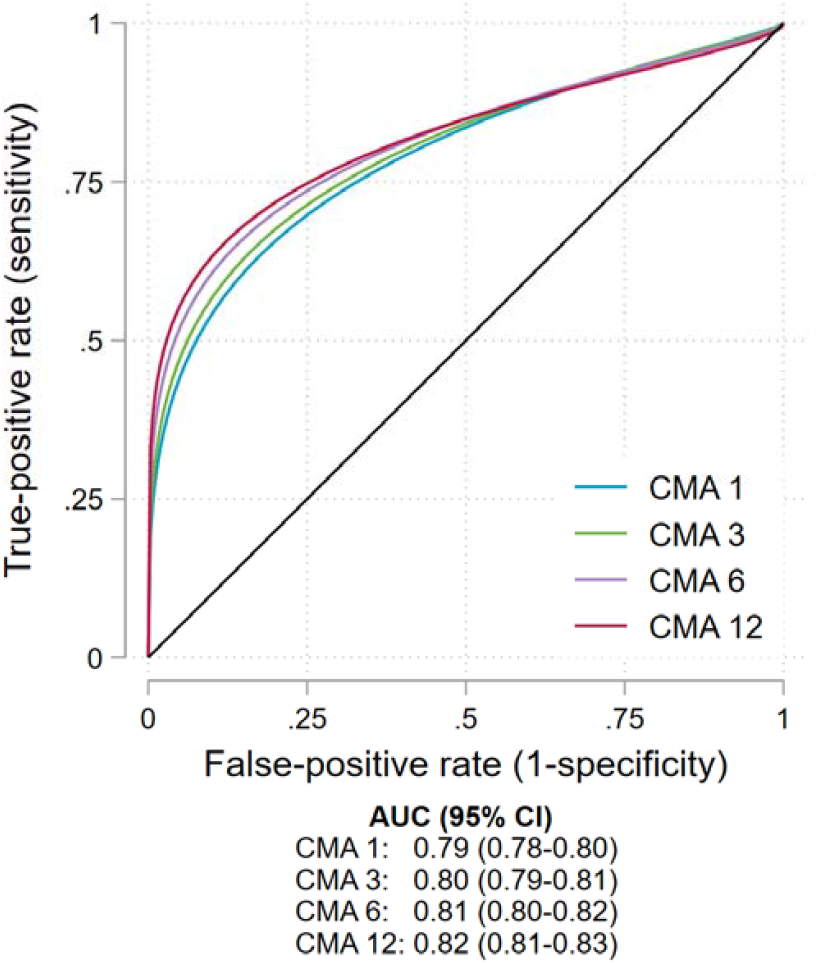
ROC curves of the accuracy of CMA for predicting viral non-suppression. True-positive rate (sensitivity), false-positive rate (1-specificity), and area under the curve (AUC) of cumulative medication availability (CMA) over 1, 3, 6, and 12 months before viral load testing for predicting viral non-suppression (VNS) at a threshold of ≥400 copies/mL. 79,463 viral load values from 28,785 participants were included in the analysis.

**Figure S3.**
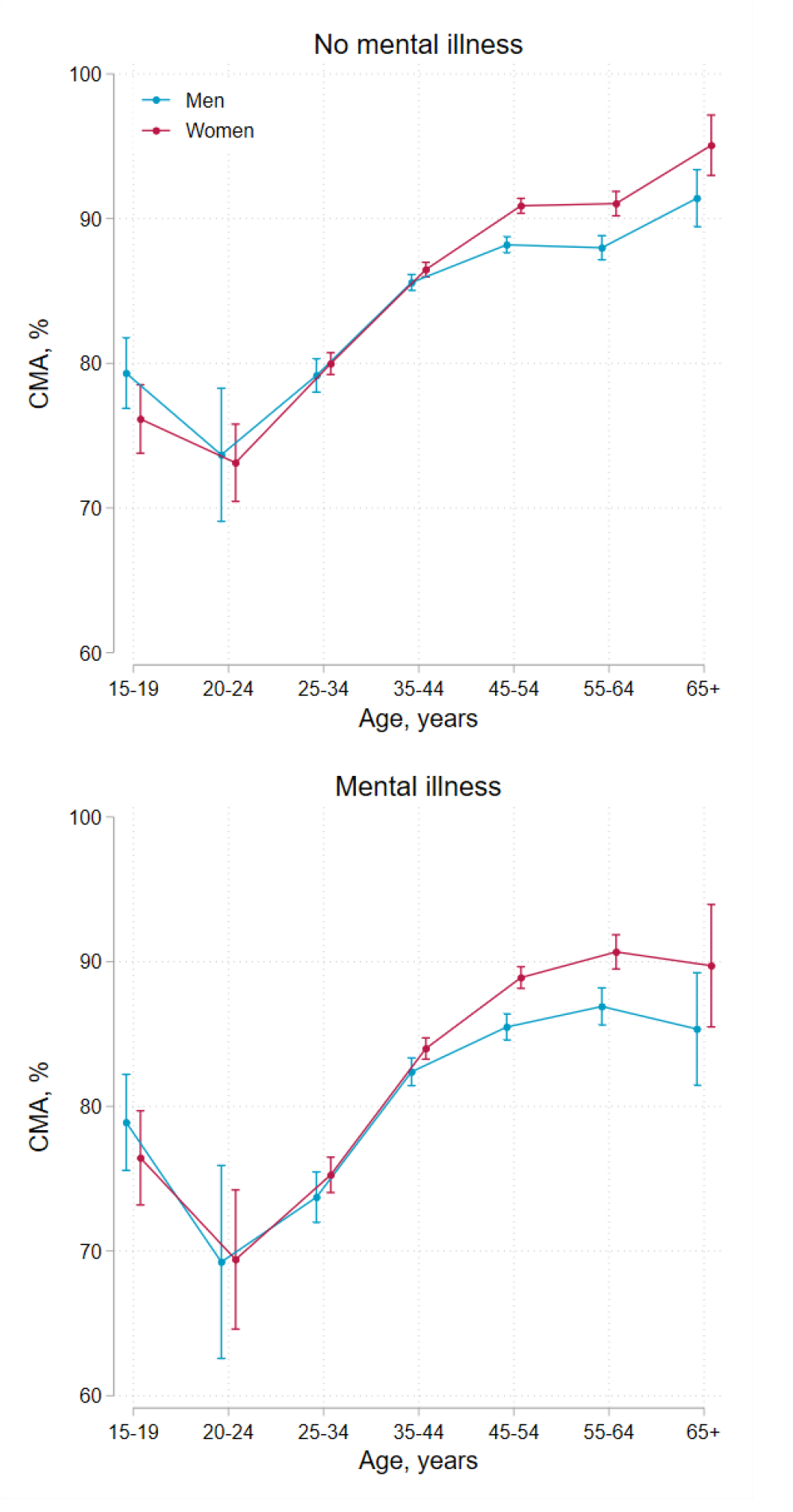
Cumulative medication availability (CMA) in the second year after baseline comparing men and women by mental health diagnosis and age group. Error bars represent 95% confidence intervals for means and proportions. N=48,645

**Figure S4.**
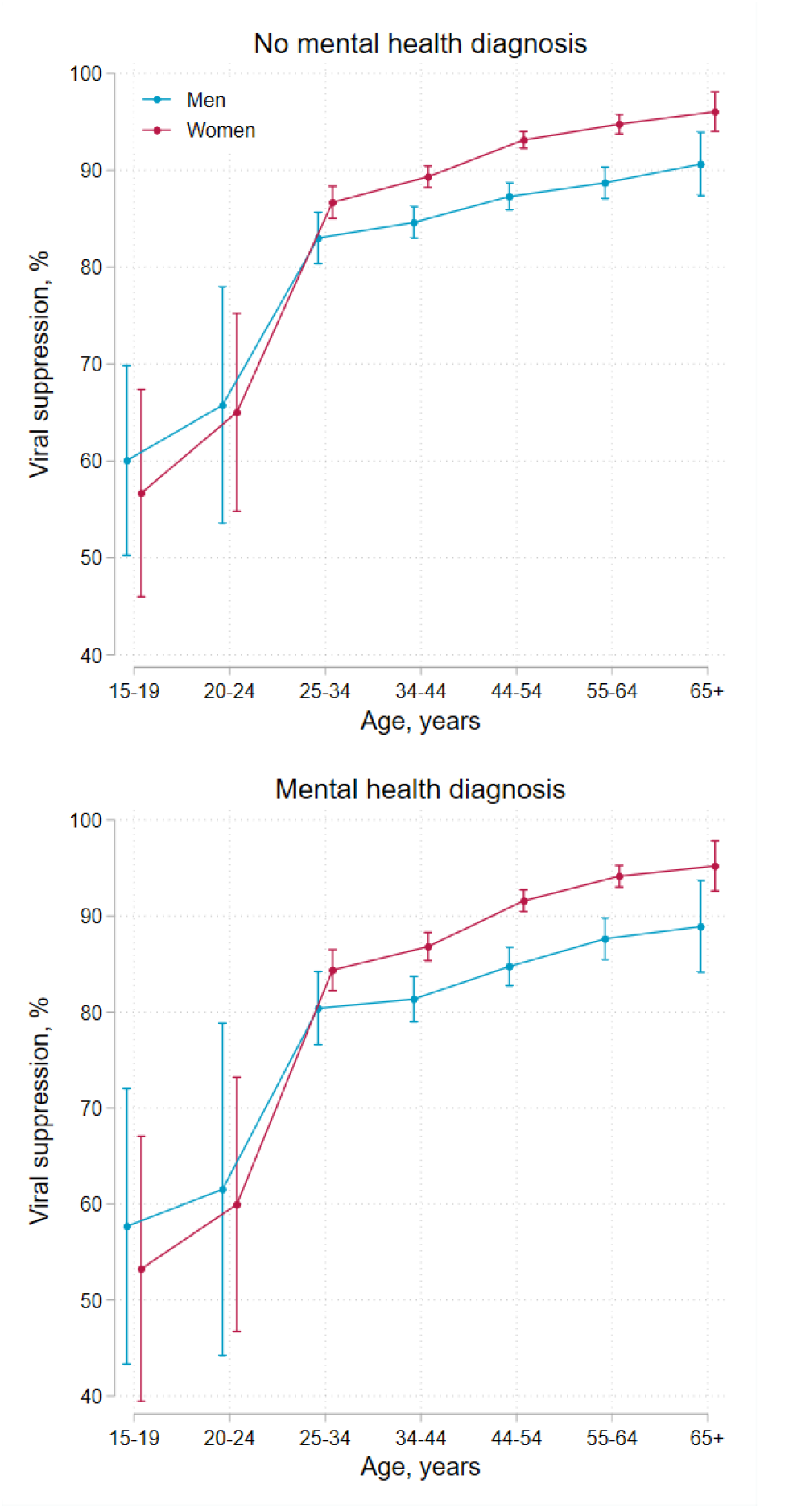
Viral suppression (viral load <400 copies/mL) at 2 years after baseline comparing men and women by mental health diagnosis and age group. Error bars represent 95% confidence intervals for means and proportions. N=28’785

**Table S1:**
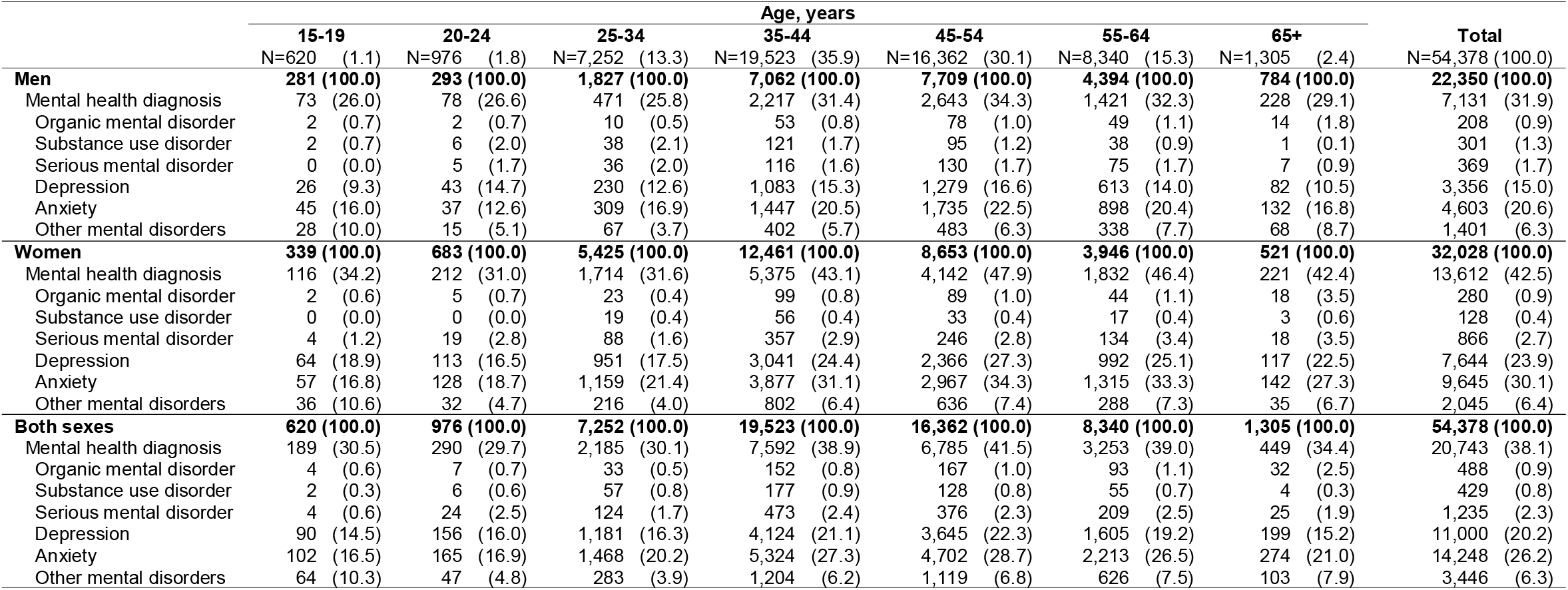
Prevalence of mental health diagnoses at the end of follow-up by sex and age at the end of follow-up.

**Table S2:** Predicted probabilities of being in each of the four adherence groups by sex, age, and mental health diagnoses at baseline.

| Men, % (95 confidence intervals) |  |  |  |  |  |  |  |  |  |  |  |  |  |  |
| --- | --- | --- | --- | --- | --- | --- | --- | --- | --- | --- | --- | --- | --- | --- |
|  | No mental illness |  |  |  |  |  |  | Mental illness |  |  |  |  |  |  |
|  | 15-19 years | 20-24 years | 25-34 years | 35-44 years | 45-54 years | 55-64 years | 65+ years | 15-19 years | 20-24 years | 25-34 years | 35-44 years | 45-54 years | 55-64 years | 65+ years |
| Continuous non-adherence | 6.8%<br>(3.9-9.6) | 21.9%<br>(12.0-31.9) | 11.2%<br>(9.9-12.5) | 5.2%<br>(4.6-5.8) | 4.7%<br>(4.0-5.3) | 4.9%<br>(3.8-6.1) | 4.8%<br>(1.0-8.6) | 11.2%<br>(6.6-15.8) | 32.7%<br>(19.7-45.7) | 18.5%<br>(16.1-20.9) | 9.1%<br>(7.9-10.4) | 8.3%<br>(7.1-9.6) | 8.8%<br>(6.7-10.9) | 8.9%<br>(2.2-15.6) |
| Increasing adherence | 10.6%<br>(7.0-14.3) | 9.2%<br>(2.2-16.1) | 7.0%<br>(5.9-8.1) | 6.9%<br>(6.2-7.5) | 5.5%<br>(4.8-6.2) | 5.8%<br>(4.5-7.1) | 3.3%<br>(0.1-6.5) | 13.8%<br>(9.1-18.5) | 10.7%<br>(2.6-18.9) | 9.0%<br>(7.5-10.6) | 9.4%<br>(8.2-10.7) | 7.8%<br>(6.6-9.0) | 8.2%<br>(6.2-10.1) | 4.8%<br>(0.2-9.4) |
| Decreasing adherence | 22.1%<br>(17.2-27.0) | 18.6%<br>(9.1-28.2) | 17.1%<br>(15.5-18.7) | 14.4%<br>(13.5-15.4) | 11.4%<br>(10.5-12.4) | 12.0%<br>(10.2-13.8) | 5.0%<br>(1.1-8.9) | 25.5%<br>(19.8-31.1) | 19.4%<br>(9.4-29.4) | 19.7%<br>(17.5-21.9) | 17.6%<br>(16.0-19.2) | 14.3%<br>(12.8-15.8) | 14.9%<br>(12.5-17.3) | 6.5%<br>(1.5-11.4) |
| Continuous high adherent | 60.5%<br>(54.7-66.3) | 50.3%<br>(38.0-62.6) | 64.7%<br>(62.6-66.7) | 73.5%<br>(72.3-74.7) | 78.4%<br>(77.1-79.6) | 77.3%<br>(75.0-79.6) | 86.9%<br>(80.9-92.9) | 49.5%<br>(43.3-55.8) | 37.2%<br>(25.5-48.8) | 52.8%<br>(50.1-55.5) | 63.8%<br>(61.8-65.8) | 69.6%<br>(67.5-71.6) | 68.2%<br>(65.0-71.3) | 79.8%<br>(71.2-88.5) |
| Women, % (95 confidence intervals) |  |  |  |  |  |  |  |  |  |  |  |  |  |  |
|  | No mental illness |  |  |  |  |  |  | Mental illness |  |  |  |  |  |  |
|  | 15-19 years | 20-24 years | 25-34 years | 35-44 years | 45-54 years | 55-64 years | 65+ years | 15-19 years | 20-24 years | 25-34 years | 35-44 years | 45-54 years | 55-64 years | 65+ years |
| Continuous non-adherence | 10.3%<br>(7.1-13.5) | 16.7%<br>(13.0-20.4) | 11.4%<br>(10.6-12.2) | 5.5%<br>(4.9-6.0) | 3.0%<br>(2.5-3.5) | 2.6%<br>(1.6-3.5) | 3.7%<br>(0.1-7.3) | 16.3%<br>(11.4-21.1) | 25.5%<br>(20.2-30.9) | 18.9%<br>(17.2-20.7) | 9.7%<br>(8.6-10.8) | 5.5%<br>(4.5-6.5) | 4.8%<br>(3.0-6.6) | 7.1%<br>(0.5-13.7) |
| Increasing adherence | 12.3%<br>(8.8-15.8) | 14.0%<br>(10.5-17.5) | 8.0%<br>(7.3-8.7) | 5.2%<br>(4.7-5.7) | 3.6%<br>(3.1-4.2) | 3.4%<br>(2.3-4.6) | 1.0%<br>(-0.9-2.9) | 15.2%<br>(10.9-19.6) | 16.8%<br>(12.5-21.1) | 10.4%<br>(9.1-11.6) | 7.4%<br>(6.4-8.3) | 5.3%<br>(4.4-6.2) | 5.0%<br>(3.3-6.7) | 1.4%<br>(0.0-4.2) |
| Decreasing adherence | 25.7%<br>(21.0-30.5) | 17.4%<br>(13.6-21.2) | 13.7%<br>(12.9-14.6) | 10.9%<br>(10.2-11.6) | 9.0%<br>(8.2-9.9) | 9.0%<br>(7.2-10.8) | 5.9%<br>(1.3-10.5) | 28.3%<br>(23.0-33.5) | 18.5%<br>(14.3-22.7) | 15.9%<br>(14.5-17.3) | 13.6%<br>(12.4-14.8) | 11.7%<br>(10.4-13.0) | 11.7%<br>(9.3-14.2) | 7.8%<br>(1.8-13.7) |
| Continuous high adherent | 51.6%<br>(46.2-57.0) | 51.9%<br>(46.8-56.9) | 66.8%<br>(65.6-68.0) | 78.4%<br>(77.4-79.3) | 84.4%<br>(83.3-85.5) | 85.0%<br>(82.7-87.3) | 89.4%<br>(83.5-95.4) | 40.2%<br>(34.8-45.7) | 39.2%<br>(34.1-44.3) | 54.8%<br>(52.8-56.8) | 69.3%<br>(67.6-71.0) | 77.5%<br>(75.8-79.3) | 78.5%<br>(75.3-81.7) | 83.7%<br>(75.0-92.4) |

**Table S3:** Viral suppression rates by sex, age group, and mental health status at 2 years after.

| Sex | Age group, years | Viral load <400 copies/mL, % (95% CI) |  | Viral load <100 copies/mL, % (95% CI) |  | Viral load <1000 copies/mL, % (95% CI) |  |
| --- | --- | --- | --- | --- | --- | --- | --- |
|  |  | No mental health diagnosis | Mental health diagnosis | No mental health diagnosis | Mental health diagnosis | No mental health diagnosis | Mental health diagnosis |
| Men | 15-19 | 60.1% (50.3-69.8) | 57.7% (43.3-72.0) | 56.1% (47.1-65.1) | 56.3% (44.1-68.5) | 66.6% (57.5-75.8) | 64.6% (51.8-77.5) |
| Men | 20-24 | 65.8% (53.6-77.9) | 61.5% (44.2-78.8) | 58.7% (46.7-70.6) | 58.4% (42.6-74.1) | 67.8% (55.8-79.7) | 66.1% (49.8-82.4) |
| Men | 25-34 | 83.0% (80.4-85.7) | 80.4% (76.6-84.2) | 80.3% (77.7-82.8) | 78.7% (75.3-82.2) | 84.5% (82.0-87.0) | 81.7% (78.1-85.4) |
| Men | 35-44 | 84.6% (83.0-86.2) | 81.3% (79.0-83.7) | 81.1% (79.5-82.6) | 78.8% (76.8-80.8) | 85.7% (84.2-87.2) | 83.2% (81.2-85.2) |
| Men | 45-54 | 87.3% (85.9-88.7) | 84.7% (82.7-86.7) | 83.7% (82.3-85.0) | 81.1% (79.3-82.9) | 88.8% (87.6-90.1) | 86.8% (85.2-88.5) |
| Men | 55-64 | 88.7% (87.1-90.3) | 87.6% (85.5-89.8) | 84.2% (82.5-85.9) | 83.6% (81.4-85.7) | 90.2% (88.7-91.7) | 89.9% (88.1-91.7) |
| Men | 65+ | 90.6% (87.4-93.9) | 88.9% (84.1-93.7) | 87.3% (83.8-90.8) | 86.1% (81.3-90.8) | 92.2% (89.3-95.2) | 90.6% (86.2-94.9) |
| Women | 15-19 | 56.7% (46.0-67.4) | 53.2% (39.4-67.1) | 48.6% (38.7-58.5) | 46.4% (33.9-59.0) | 61.3% (51.3-71.3) | 58.3% (45.5-71.1) |
| Women | 20-24 | 65.0% (54.8-75.2) | 60.0% (46.7-73.2) | 62.6% (53.4-71.8) | 60.6% (49.4-71.8) | 70.7% (61.0-80.3) | 68.6% (57.1-80.1) |
| Women | 25-34 | 86.7% (85.0-88.3) | 84.4% (82.2-86.5) | 84.2% (82.6-85.8) | 82.2% (80.2-84.1) | 88.0% (86.5-89.6) | 85.7% (83.7-87.6) |
| Women | 35-44 | 89.3% (88.2-90.4) | 86.8% (85.3-88.3) | 87.0% (85.9-88.0) | 84.8% (83.5-86.0) | 90.6% (89.6-91.5) | 88.8% (87.6-89.9) |
| Women | 45-54 | 93.1% (92.2-94.0) | 91.6% (90.4-92.7) | 90.8% (90.0-91.7) | 88.9% (87.8-90.0) | 94.2% (93.5-95.0) | 93.1% (92.2-94.0) |
| Women | 55-64 | 94.7% (93.7-95.7) | 94.1% (93.0-95.3) | 92.1% (91.0-93.3) | 91.5% (90.2-92.7) | 95.5% (94.6-96.4) | 95.3% (94.3-96.2) |
| Women | 65+ | 96.0% (94.0-98.1) | 95.2% (92.6-97.8) | 92.9% (90.2-95.6) | 91.8% (88.5-95.1) | 96.9% (95.2-98.6) | 96.2% (93.8-98.5) |

Abbreviations: CI=confidence interval

**Table S4:** Sensitivity analysis of associations between mental health diagnoses and non-adherence.

|  | Primary analysis | Sensitivity analyses |  |  |
| --- | --- | --- | --- | --- |
|  | CMA <80%<br>aRR (95% CI) | CMA <70%<br>aRR (95% CI) | CMA <90%<br>aRR (95% CI) | CMA <80%<br>Repeated mental<br>health diagnoses,<br>aRR (95% CI) |
| Mental health diagnosis |  |  |  |  |
| Organic mental disorder | 1.31 (1.12-1.54) | 1.49 (1.24-1.78) | 1.21 (1.06-1.38) | 1.30 (0.98-1.71) |
| Substance use disorder | 1.60 (1.41-1.83) | 1.66 (1.41-1.94) | 1.51 (1.36-1.68) | 1.60 (1.28-1.99) |
| Serious mental disorder | 1.23 (1.11-1.36) | 1.26 (1.12-1.42) | 1.19 (1.09-1.29) | 1.12 (0.97-1.29) |
| Depression | 1.21 (1.17-1.25) | 1.23 (1.17-1.28) | 1.17 (1.14-1.21) | 1.20 (1.15-1.25) |
| Anxiety | 1.22 (1.18-1.26) | 1.23 (1.19-1.28) | 1.19 (1.16-1.22) | 1.19 (1.15-1.24) |
| Other mental health diagnoses | 1.10 (1.03-1.17) | 1.13 (1.05-1.21) | 1.07 (1.02-1.13) | 0.96 (0.86-1.07) |
| Any mental health diagnosis | 1.21 (1.18-1.25) | 1.23 (1.19-1.28) | 1.18 (1.15-1.20) | 1.18 (1.14-1.22) |

Risk ratios were adjusted for years since baseline, age group, and sex.
Abbreviations: CMA=cumulative medication availability, aRR=adjusted risk ratio, CI=confidence interval

**Table S5:** Sensitivity analysis of associations between mental health diagnoses and viral non-suppression.

|  | Primary analysis | Sensitivity analyses |  |  |
| --- | --- | --- | --- | --- |
|  | Viral load >400<br>copies/mL<br>aRR (95% CI) | Viral load >100<br>copies/mL<br>aRR (95% CI) | Viral load >1000<br>copies/mL<br>aRR (95% CI) | Viral load >400 copies/mL<br>Repeated mental health<br>diagnoses<br>aRR (95% CI) |
| Mental health diagnosis |  |  |  |  |
| Organic mental disorder | 1.78 (1.41-2.24) | 1.74 (1.45-2.09) | 1.74 (1.35-2.25) | 1.90 (1.30-2.77) |
| Substance use disorder | 1.82 (1.42-2.34) | 1.69 (1.37-2.09) | 1.88 (1.44-2.46) | 2.06 (1.38-3.08) |
| Serious mental disorder | 1.54 (1.30-1.82) | 1.48 (1.29-1.71) | 1.62 (1.36-1.94) | 1.53 (1.23-1.90) |
| Depression | 1.26 (1.17-1.35) | 1.23 (1.16-1.30) | 1.29 (1.20-1.39) | 1.30 (1.20-1.41) |
| Anxiety | 1.11 (1.04-1.19) | 1.08 (1.02-1.14) | 1.10 (1.03-1.18) | 1.14 (1.05-1.23) |
| Other mental health diagnoses | 1.10 (0.98-1.23) | 1.07 (0.98-1.18) | 1.05 (0.93-1.19) | 1.01 (0.84-1.21) |
| Any mental health diagnosis | 1.16 (1.09-1.24) | 1.14 (1.08-1.20) | 1.17 (1.09-1.24) | 1.21 (1.13-1.30) |

RRs were adjusted for years since baseline, age group and sex.
Abbreviations: aRR=adjusted risk ratio, CI=confidence interval

**Text S1: Detailed description of statistical methods to model non-adherence and viral non-suppression**

To model non-adherence, we split patients’ follow-up time into consecutive 12-month intervals, estimated the mean adherence during each interval, and dichotomised continuous adherence values at the prespecified non-adherence thresholds (e.g. 80%). The analysis dataset contained one binary outcome for non-adherence for each patient and each completed year of follow-up. Using this dataset, we estimated adjusted risk ratios for factors associated with non-adherence using modified mixed-effects Poisson regression models with robust standard errors and a random intercept at patient level. First, we fitted seven adjusted models to estimate risk ratios for each of the six groups of mental health diagnoses and any mental disorder adjusting for years since baseline, age, sex, and an interaction term between age and sex. Next, we fitted a model including an indicator for each of the six groups of mental health diagnoses, years since baseline, age, sex, and an interaction term between age and sex. We used contrasts to estimate differences in the risk of non-adherence between age groups, men and women, and patients with and without mental health diagnoses. Poisson regression models were used to overcome issues with the convergence of log-binomial models. We dichotomised adherence scores because data were heavily left-skewed.

In analysis of factors associated with viral non-suppression, the dataset contained one binary outcome for each patient and viral load test result. We fitted the models described above to estimate adjusted risk ratios for factors associated with viral non-suppression. In addition, we fitted seven models adjusting risk ratios for each group of mental health diagnoses for cumulative medication availability, age, sex, and year since baseline. We estimated differences in the risk of viral non-suppression between age groups, men and women, and patients with and without mental health diagnoses using contrasts. Finally, we estimated and plotted the probability of viral suppression (viral load <400 copies) at 2 years after baseline for people with and without mental health diagnoses by age and sex using predictive margins and a model including binary indicators for any mental disorder and sex, categorical variables for years since baseline, age, and interaction terms between age and sex, any mental disorder and sex and any mental disorder and age.

